# Computed tomography radiomics-based cross-sectional detection of mandibular osteoradionecrosis in head and neck cancer survivors

**DOI:** 10.1101/2024.09.11.24313485

**Authors:** MD Anderson Head and Neck Cancer Symptom Working Group, Serageldin Kamel, Laia Humbert-Vidan, Zaphanlene Kaffey, Abdulrahman Abusaif, David T. A. Fuentes, Kareem Wahid, Cem Dede, Mohamed A. Naser, Renjie He, Ahmed W. Moawad, Khaled M. Elsayes, Melissa M. Chen, Adegbenga O. Otun, Jillian Rigert, Mark Chambers, Andrew Hope, Erin Watson, Kristy K. Brock, Katherine Hutcheson, Lisanne van Dijk, Amy C. Moreno, Stephen Y. Lai, Clifton D. Fuller, Abdallah S. R. Mohamed

## Abstract

**Purpose:** This study aims to identify radiomic features extracted from contrast-enhanced CT scans that differentiate osteoradionecrosis (ORN) from normal mandibular bone in patients with head and neck cancer (HNC) treated with radiotherapy (RT).

**Materials and Methods:** Contrast-enhanced CT (CECT) images were collected for 150 patients (80% train, 20% test) with confirmed ORN diagnosis at The University of Texas MD Anderson Cancer Center between 2008 and 2018. Using PyRadiomics, radiomic features were extracted from manually segmented ORN regions and the corresponding automated control regions, the later defined as the contralateral healthy mandible region. A subset of pre-selected features was obtained based on correlation analysis (r > 0.95) and used to train a Random Forest (RF) classifier with Recursive Feature Elimination. Model explainability SHapley Additive exPlanations (SHAP) analysis was performed on the 20 most important features identified by the trained RF classifier.

**Results:** From a total of 1316 radiomic features extracted, 810 features were excluded due to high collinearity. From a set of 506 pre-selected radiomic features, the optimal subset resulting on the best discriminative accuracy of the RF classifier consisted of 67 features. The RF classifier was well calibrated (Log Loss 0.296, ECE 0.125) and achieved an accuracy of 88% and a ROC AUC of 0.96. The SHAP analysis revealed that higher values of Wavelet-LLH First-order Mean and Median were associated with ORN of the jaw (ORNJ). Conversely, higher Exponential GLDM Dependence Entropy and lower Square First-order Kurtosis were more characteristic of normal mandibular tissue.

**Conclusion:** This study successfully developed a CECT-based radiomics model for differentiating ORNJ from healthy mandibular tissue in HNC patients after RT. Future work will focus on the detection of subclinical ORNJ regions to guide earlier interventions.

## Introduction

Osteoradionecrosis of the jaw (ORNJ) is a debilitating long-term complication of radiotherapy (RT) that significantly impacts the quality of life of survivors of HNC, especially oropharyngeal cancer (OPC). Despite reductions in smoking, the incidence of OPC cancer is on the rise due to the increasing incidence of human papillomavirus (HPV)-associated OPC ^1^. It is projected that hundreds of thousands of locally advanced OPC patients will receive radiation as a primary treatment modality with an expected high rate of long-term survivors ^2^. These patients face an increased cumulative lifetime risk for the development of long-term complications from RT, such as ORNJ. Early diagnosis and intervention are crucial for improving outcomes, particularly for high-risk patients where mandibular RT dose constraints cannot be achieved due to tumor proximity. Currently, clinicians utilize basic preventive measures to reduce the risk of ORNJ, such as removing compromised teeth before radiation therapy and avoiding dental extractions within radiation fields after radiation treatment ^3^. However, the development and implementation of models that identify subclinical mandibular subvolumes at high risk for ORNJ could inform guidelines for preventative and conservative interventions aimed to reduce the incidence and severity of ORNJ. Recently, predictive models that utilize radiological imaging features of normal tissue have been validated for accurately forecasting various RT-induced complications such as xerostomia and pneumonitis ^4,5^.

Radiomics is a medical image analysis method involving high-throughput extraction of quantitative features from 2- or 3-dimensional images. By converting digital images into mineable high-dimensional data, radiomic features give means to describe the texture, shape, and size of a defined region of interest (ROI), which can reveal characteristics imperceptible to the naked eye. These features have been shown to be valuable in predicting different therapy outcomes such as survival, local control, toxicities ^4,5^, and detection or assessment of various pathologies. ^6–17^

In this study, we aim to explore the utility of radiomic feature analysis on the standard-of-care contrast-enhanced computed tomography (CECT) scans to distinguish mandibular ORNJ from normal mandibular bone in patients who underwent definitive radiotherapy for head and neck cancer. Our work represents a crucial step towards developing a model for the early detection of ORNJ changes in the mandible, prior to conventional radiologic or clinical diagnosis. Additionally, we aim to evaluate the most effective statistical models for the binary classification of ORNJ versus normal bone based on the identified radiomic features.

## Materials and methods

### Study Design

This retrospective study was approved by the institutional review board (IRB) and the prerequisite for informed consent was waived [RCR-03-800]. Patients treated at the University of Texas MD Anderson Cancer Center were identified by querying the radiology reports of head and neck CT studies conducted between 2008 to 2018 using mPower Clinical Analytics (Nuance Communications Inc., Burlington, MA). The Boolean search strategy of the following keywords: “Osteoradionecrosis” AND “Squamous” AND (“Radiation” OR “Radiotherapy” OR “IMRT”) resulted in an initial pool of 676 patients. Post-radiotherapy CECTs were directly retrieved from the institutional picture and archiving communication system (PACS) for manual screening and verification of findings consistent with ORNJ by a second observer. The earliest CECT with verified radiographic findings of ORNJ, which included bone sequestration, cortical bone erosion, a mix of sclerotic and osteolytic changes, soft tissue involvement with thickening or abscesses, and/or pathological fractures was included. Patients were excluded if they had irrelevant diagnoses (N=237), bilateral ORNJ at comparable sites (e.g., at both mandibular rami) [N=8], non-mandibular ORN (N=78), or history of major mandibular surgeries (N=203). No metal artifacts were present in the ORNJ or contralateral control ROIs in the final cohort. After exclusions, a final set of 150 patients were analyzed (Figure 1). The primary endpoint was physician-reported diagnosis of ORNJ of any grade. ^18^

**Figure 1.**
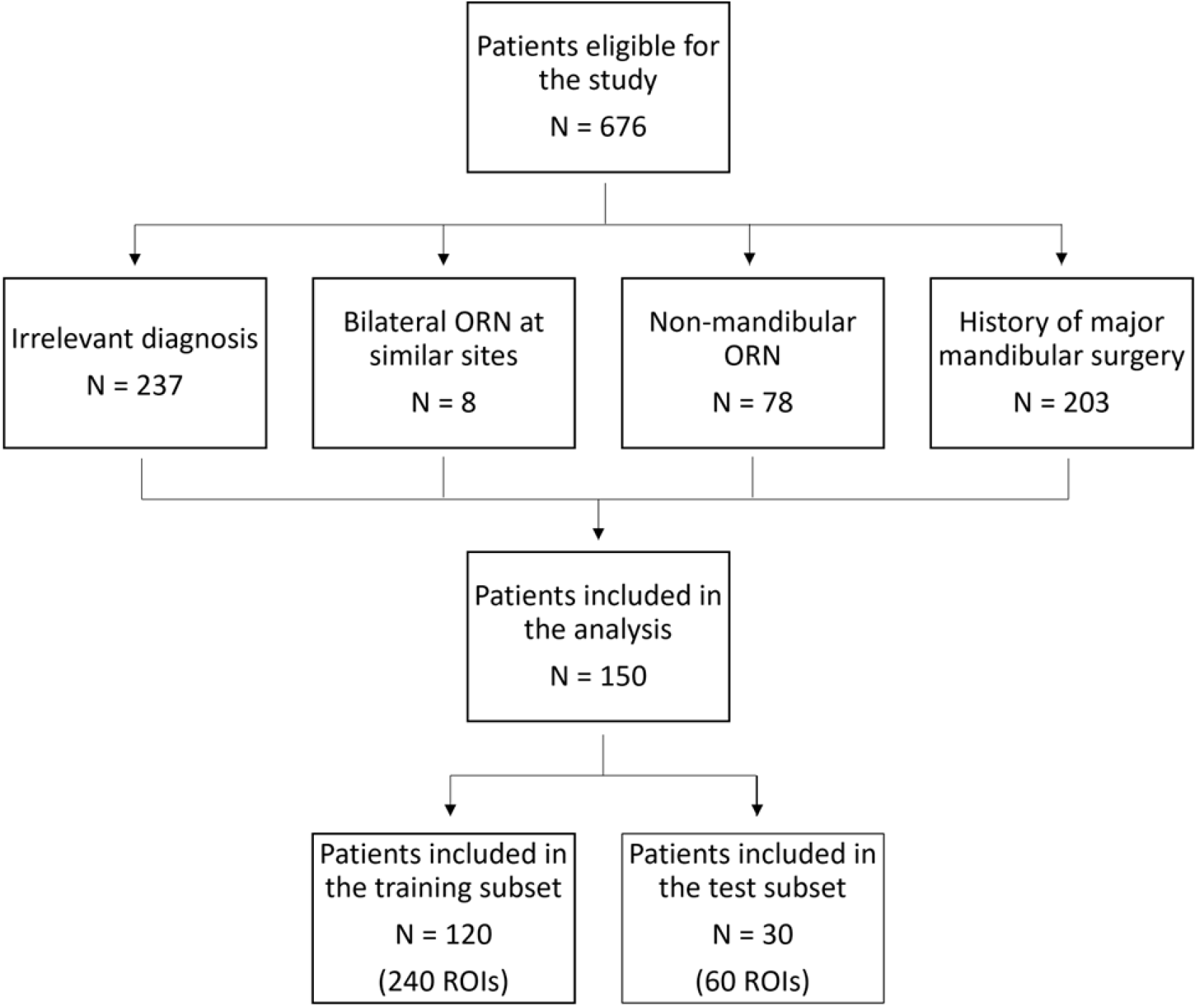
Patient exclusion and dataset split workflow.

### Image Acquisition

All CT scans were acquired with a multi-detector row CT scanner (Lightspeed 16, GE Healthcare, Milwaukee, WI) according to our institution’s protocol. The acquisition parameters ranged from 1-3 mm section thickness, with a median of 1 mm. The X-ray tube current ranged from 99-584 mA at a peak voltage between 120-140 kVp. Composition of all images was 512 × 512 pixels. The post-intravenous contrast phase CT series were acquired at a 90-second delay following the injection of 120 CC of contrast agent. All selected Digital Imaging and Communications in Medicine (DICOM) images were converted to Neuroimaging Informatics Technology Initiative NIfTI format.

### Image Segmentation

The mandibular volumes affected by ORNJ were manually segmented by a trained research fellow (A.A.) using Amira Software (Thermo Scientific, Waltham, MA, USA) and reviewed by a radiation oncologist (A.M.) with 10 years of experience. In order to isolate ORN-specific radiomic features, a conservative segmentation approach was adopted to avoid the inclusion of adjacent healthy bone tissue. Following ORNJ segmentations, mirroring “control” volumes representing healthy tissue were created on the contralateral mandible free of ORN (Figure 2). Thus, for each subject in the dataset, an ORNJ and a control ROI were obtained. We utilized the Convert3D image-processing tool of ITK-SNAP software ^19^to horizontally flip the segmented ORNJ volume around the x-axis thus defining the control ROI. The resulting control volumes were subsequently reviewed for the validity of their anatomical location. Due to discrepancies in anatomical orientation, the x-axis of the CT images did not consistently correspond to the midline of the mandible. Therefore, manual repositioning for misaligned control volumes was required to ensure accurate correspondence with their contralateral counterparts without modifications to their shape or size.

**Figure 2.**
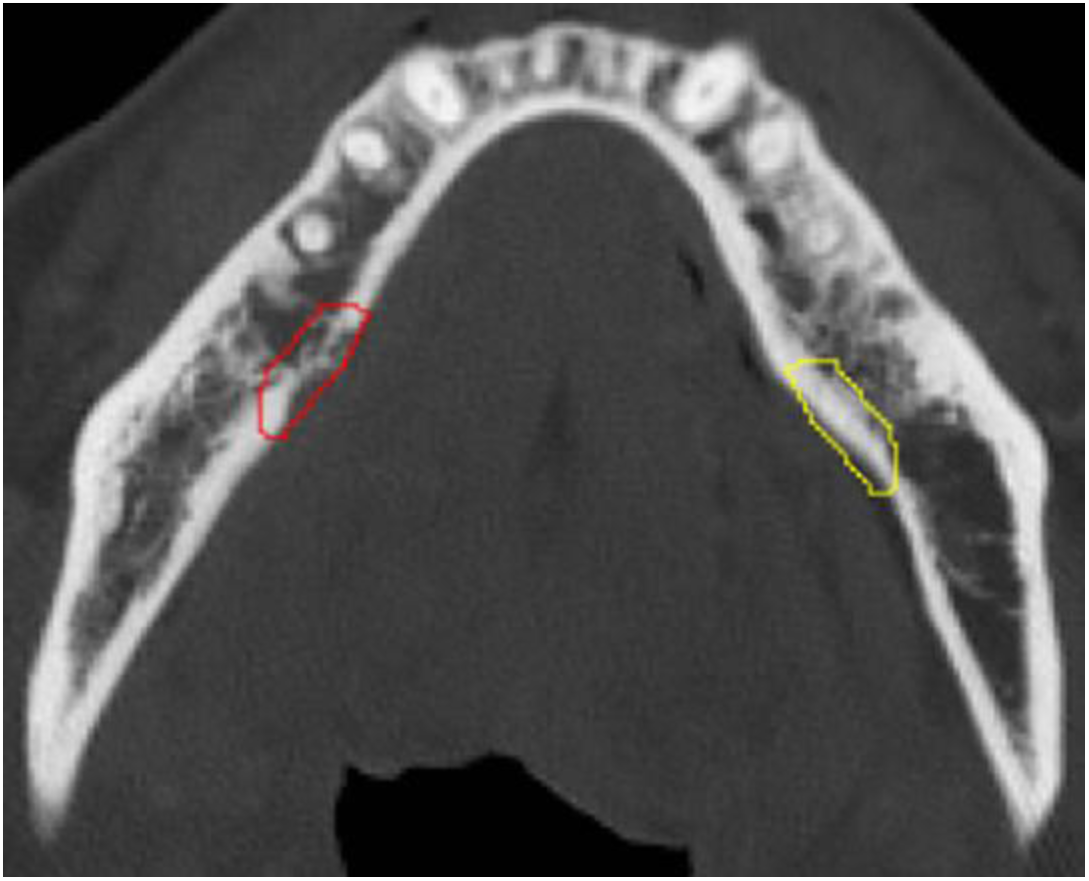
Axial view of a contrast-enhanced CT image with segmented ORN (red) and contralateral mirror-image control normal bone (yellow).

### Radiomic Features Extraction

Radiomic features from the described ROIs were extracted using PyRadiomics, an open-source Python library for high-throughput extraction of quantitative features from medical imaging ^20^. A fixed bin-width of 3.2 Hounsfield Units (HU) was used for grey value discretization with a target bin number of 30-130. The extracted features included first-order statistical features, shape-based features, and higher-order textural features, derived through matrix-based and filter-based methods ^21,22^. Additionally, an array of imaging filters was applied to CT images to diversify the radiomic features space and augment different textural and intensity characteristics. These filters included the Laplacian of Gaussian, which accentuates edges by highlighting regions of rapid intensity transition, and various wavelet transforms, decomposing the image into multiple frequency sub-bands. Furthermore, mathematical transformations such as square, square root, logarithmic, and exponential filters were applied. In total, 1316 features were used for further analysis. The detailed explanations of the radiomic features and the intrinsic filters used in PyRadiomics are listed in the platform documentation ^23^, which is in accordance with the Imaging Biomarker Standardization Initiative (IBSI) ^24^.

### Data Preparation and Feature Normalization

The dataset was split into training (80%) and testing (20%) subsets based on unique patient identifiers using a fixed random state for reproducibility. This approach ensured patient-level separation between training and testing datasets with equal distribution of ORNJ and control ROIs. The radiomic features were subsequently normalized to have a zero mean and a unit standard deviation (i.e., Gaussian-like distribution). Normalization was performed on the train subset, and the same transformation was subsequently applied to the test data to prevent information leakage. To address multicollinearity and enhance model interpretability, feature preselection was performed using a pairwise correlation analysis with a Pearson coefficient threshold of 0.95, thus reducing feature redundancy and dimensionality.

### Model Training and Evaluation

The training dataset was used for feature selection and model development. A Random Forest (RF) classifier was trained with Recursive Feature Elimination (RFE) following a 5-fold Cross-Validation (CV) approach to identify the optimal set of radiomic features (Figure 3). This approach systematically evaluates the contribution of each feature to model performance, described by predictive accuracy, iteratively removing the least contributing features. The optimal RF classifier hyperparameters, determined from a Grid Search hyperparameter tunning, were set at 100 estimators, a maximum depth of 5, a minimum of 3 samples per leaf, and ‘log2’ for the maximum features.

**Figure 3.**
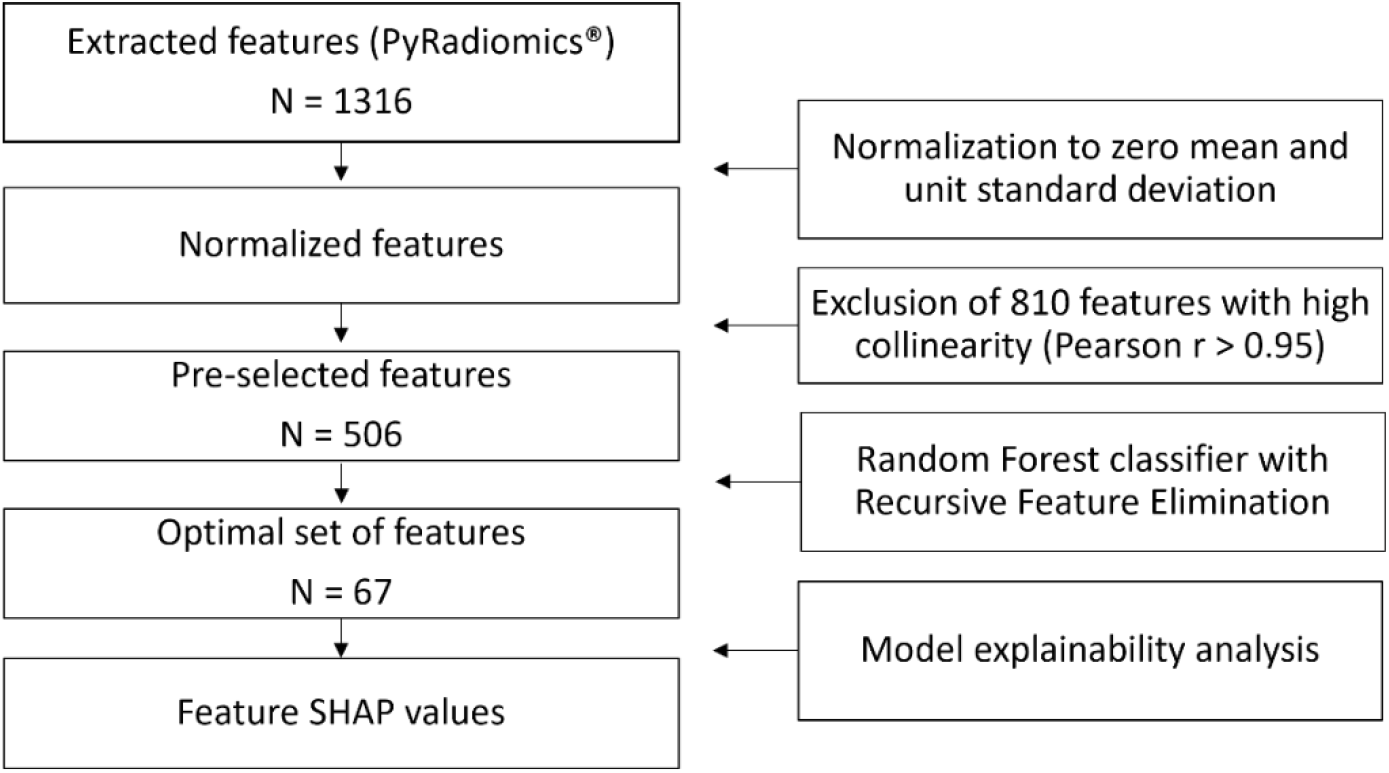
Workflow for the radiomic feature extraction and modeling steps. The number of features at each step is denoted by N.

### Model Explainability Analysis

The selected features by the RF classifier were ranked based on their importance with regards to their influence on the model performance. To explain how individual features influenced specific patient predictions, SHapley Additive exPlanations (SHAP) ^25^ values were computed. SHAP values for each feature represent a weighted average of the difference between the model’s prediction with and without a specific feature where positive, negative, or zero values indicated an increase, decrease, or no contribution to the model output, respectively.

## Results

### Patient Characteristics

A total of 150 patients with radiographic ORNJ post-RT for head and neck cancer were included in the final analysis. The mean age of the patients at the time of CECT acquisition was 62.3 years (range 27-82). The average time from the commencement of RT to the onset of ORNJ was approximately 32.6 months. The detailed breakdown of patient, disease, and treatment characteristics is provided in Appendix A.

### Radiomic Features

A total of 1316 radiomic features were initially extracted from 300 ROIs representing ORNJ (n=150) and contralateral healthy mandibles (n=150). The training set comprised 240 ROIs (120 ORNJ and 120 healthy), while the testing set consisted of 60 ROIs, equally divided between ORNJ and healthy mandibles. The correlation analysis led to the exclusion of 810 features with high collinearity (Pearson correlation coefficient > 0.95), leaving 506 features for model development. Following the RF with RFE-CV modeling process, a final set of 67 optimal features were identified (see Appendix B), where features with zero importance were discarded. The cumulative importance of the features in this final set is shown in Figure 4A. These features were ranked by their importance in the classification process of the RF model, the top 20 features are shown in Figure 4B.

**Figure 4.**
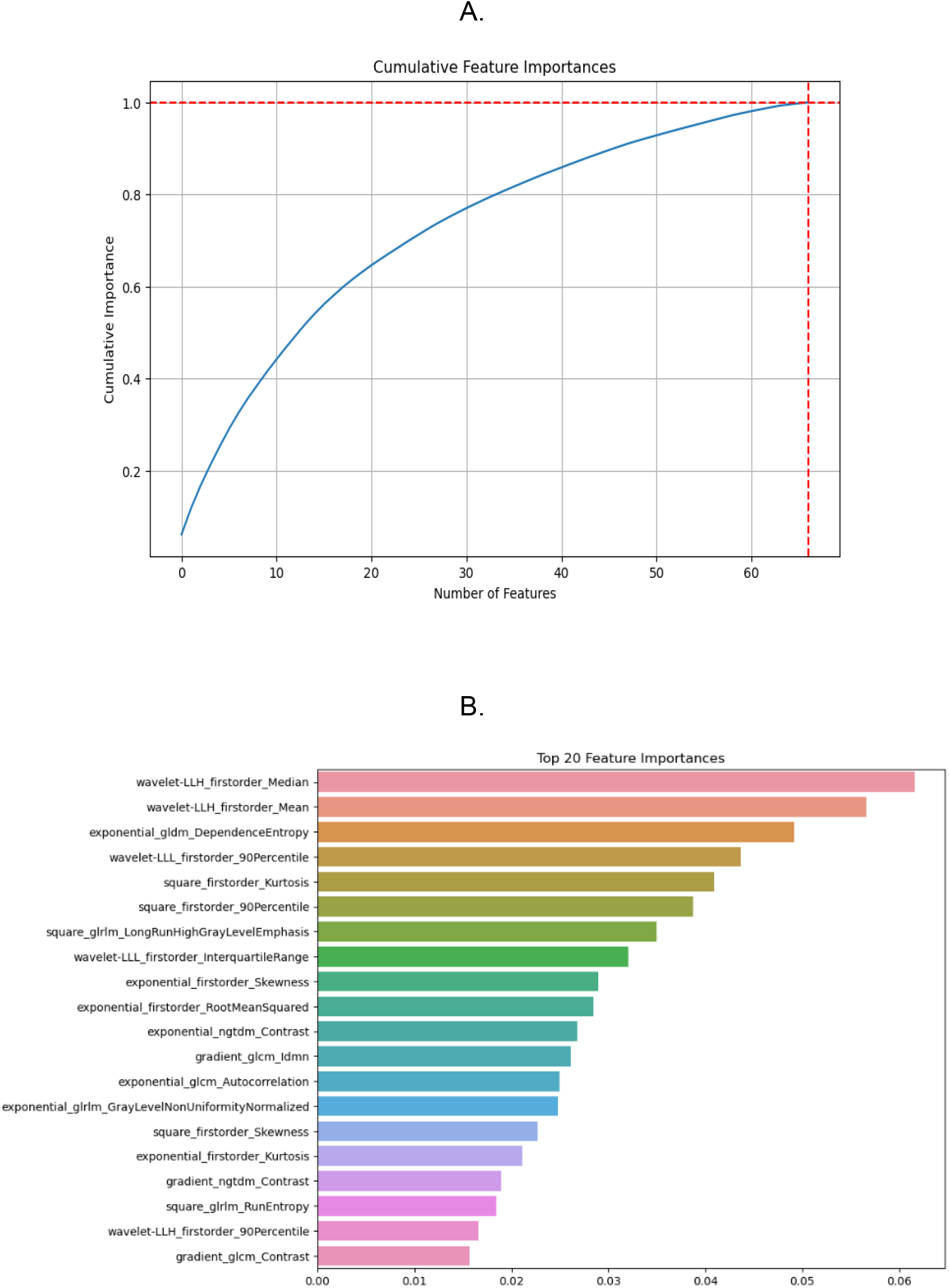
A) The cumulative sum of feature importance as determined by the Random Forest model. The horizontal line represents the threshold where cumulative importance reaches 100%, and the vertical line indicates the number of features required to achieve this cumulative importance. B) The top 20 features ranked by their importance in the classification process of the RF model. The length of each bar reflects the relative importance of each feature in the model’s decision-making process, highlighting which features are most influential in differentiating between ORNJ and healthy mandibular tissues.

### Model Performance

Model performance was assessed in terms of discrimination and calibration. The 5-fold cross-validation conducted on the training dataset resulted in an average AUC of 0.94 (range 0.90 to 0.96). For the independent test dataset, the RF model achieved an accuracy of 88% and an AUC of 0.96 (Figure 5A). The sensitivity for correctly identifying ORNJ cases was 93%, with 28 out of 30 ORNJ ROIs accurately predicted. For healthy mandibular tissue, the RF model correctly predicted 25 out of 30 cases, resulting in a specificity of 83%. The trained RF classifier was overall reasonably well calibrated (Log Loss 0.296, ECE 0.125) with a slight over and under confidence of the model’s predictions at low and high probabilities, respectively (Figure 5B).

**Figure 5.**
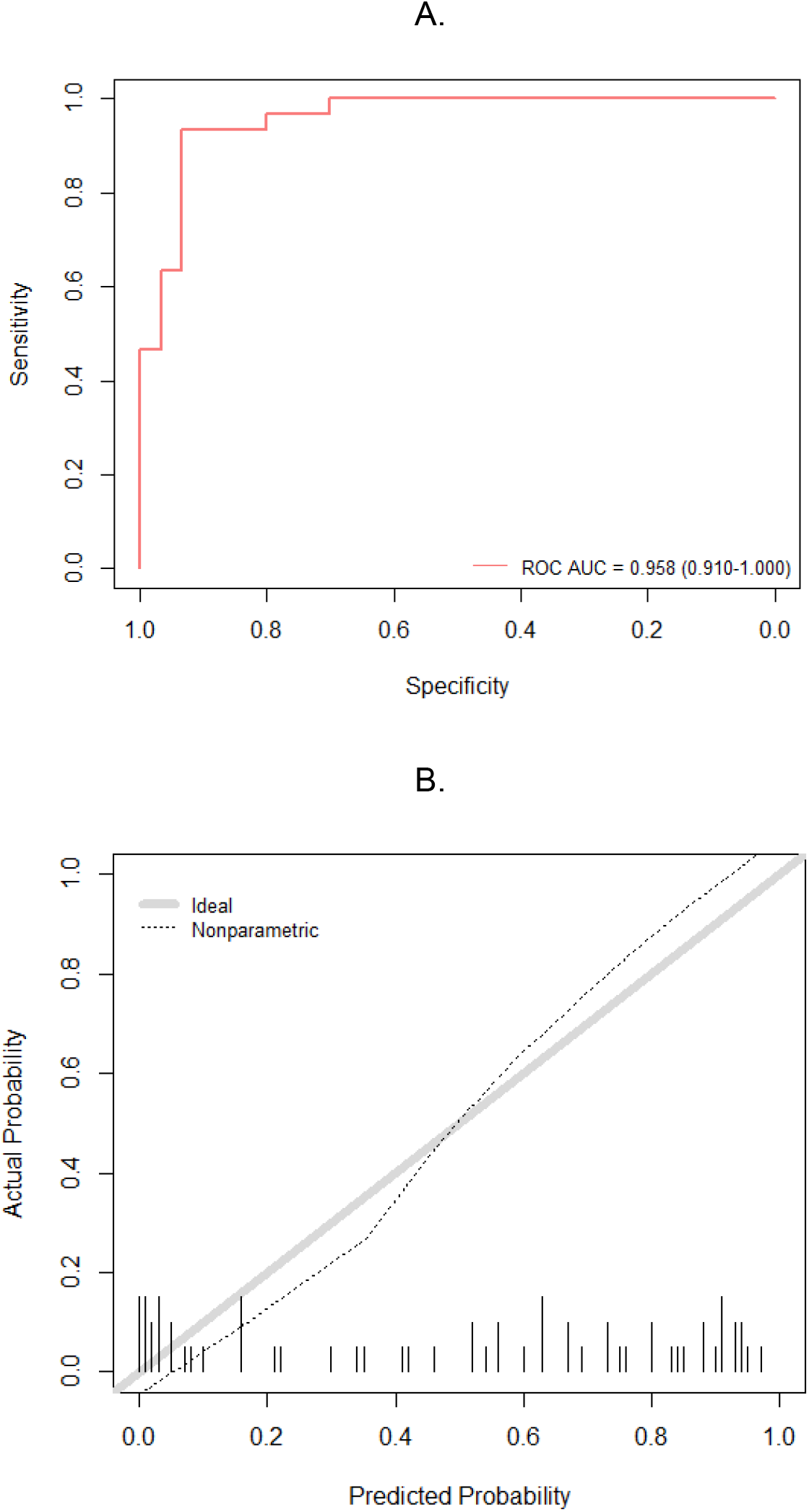
Model performance evaluation. A) Receiver operating characteristic (ROC) curves (mean and per CV fold) of the Random Forest trained on the selected extracted radiomic features (n=67) from ORN and healthy mandible VOIs on CECT. B) Reliability curve for the Random Forest classifier. Actual outcome probabilities are plotted against predicted probabilities. The thick grey diagonal line represents an ideal calibration, where predicted probabilities align perfectly with the observed outcome frequencies. Deviations from this line indicate overconfidence (points below the diagonal) or underconfidence (points above the diagonal) in the model’s predictions. The tick marks along the x-axis show the distribution of predicted probabilities. A high concentration of ticks in a certain region indicates that many predictions fall within that probability range.

### Model Explainability Analysis

As shown in Figure 6, the SHAP analysis demonstrated the strong variability of the topmost important feature values between ORNJ and normal mandibular tissues. Notably, higher values of Wavelet-LLH First-order Mean and Median, and Wavelet-LLL First-order Interquartile Range and 90th Percentile were strongly associated with ORNJ. Conversely, normal mandibular tissue was associated with higher Exponential GLDM Dependence Entropy, Wavelet-LLL First-order Interquartile Range and 90th Percentile as well as lower Square First-order Kurtosis.

**Figure 6.**
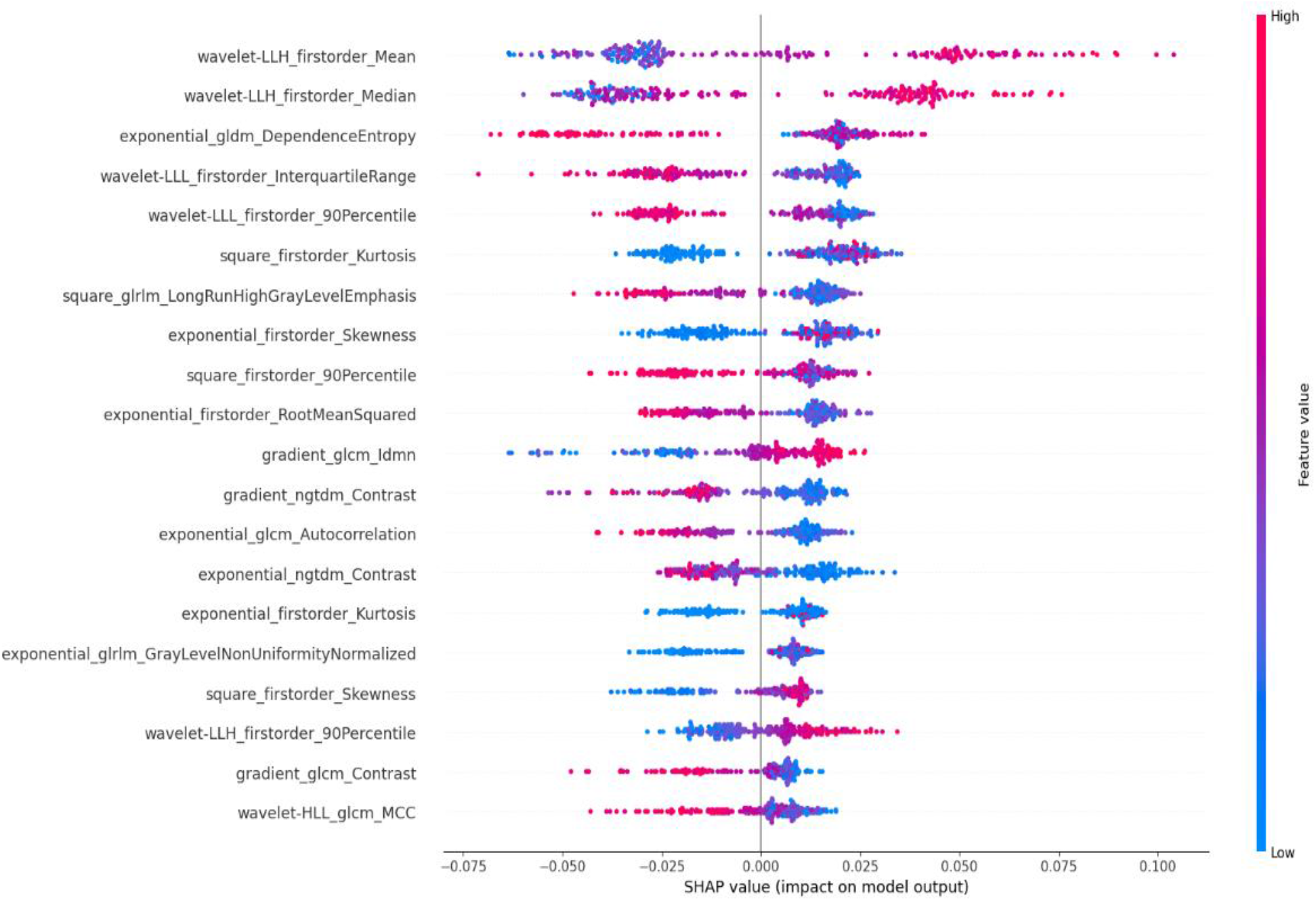
SHAP values for the top 20 most influential radiomic features. The y-axis lists each feature, ordered by the average magnitude of their SHAP values. Each dot represents the SHAP value for each individual subject in the dataset illustrating the extent of each feature’s impact on the model’s prediction for differentiating between ORNJ and healthy mandibular tissue. The color gradient, ranging from blue to red, indicates the range of feature values, with red signifying higher values.

## Discussion

The use of radiomics in oncology has been increasingly recognized for its potential in various aspects of cancer care, including predicting treatment outcomes, assessing response to therapy, and detecting early complications. However, its application in the specific context of ORNJ has been less explored. Traditional methods of diagnosing ORNJ rely heavily on clinical assessment and conventional imaging findings, which may not detect early subclinical changes. By contrast, the high-throughput analysis of radiomic features offers a more nuanced and comprehensive assessment of tissue characteristics.

In this study, we have developed and validated a CECT-based radiomics model for differentiating ORNJ from healthy mandibular tissue in head and neck cancer patients after RT. The predictive performance of the developed model comprising 67 features demonstrated significant accuracy, sensitivity, and specificity.

The SHAP analysis provided a detailed understanding of feature contributions towards the RF model’s decision-making process. The *wavelet-LLH_firstorder_Mean* and wavelet-*LLH_firstorder_Median* features, which represent the average and median pixel intensities in the LLH wavelet-transformed image, showed a positive SHAP value. This suggests that higher values of these features are strongly associated with the presence of ORN, potentially indicating regions with specific texture characteristics or density variations that are captured in this wavelet sub-band. Conversely, *gldm_DependenceEntropy*, capturing the complexity of texture in terms of exponential intensity levels and spatial dependence, demonstrated a negative SHAP value. Lower values of this feature contribute to an ORNJ classification, hinting at reduced complexity and dependence in the spatial arrangement of pixel intensities in ORNJ regions compared to controls.

Both interquartile and 90^th^ percentile first order features describing the spread of the pixel intensity values in the LLL wavelet domain, also exhibited negative SHAP values. The association of lower values of these features with ORNJ suggests that the intensity distribution within the LLL wavelet-transformed image is more homogenous or lacks extreme values in the ORNJ affected regions. These findings emphasize the importance of wavelet-based texture features in differentiating ORNJ from non-ORN tissue. The variation in the SHAP values for these features underscores the heterogeneity inherent in ORN-affected tissues on CECT images. Overall, the SHAP values provide an interpretable explanation of a relatively complex radiomic signature of ORNJ compared to the healthy mandible. The quantitative evidence supports the potential utility of CECT-based radiomic profiling in the non-invasive assessment and possible early detection of ORNJ post-RT in head and neck cancer patients.

Several studies have validated binary classification models to predict normal tissue complications of RT based on CECT texture analysis alone. For example, van Dijk et al. [4] analyzed the CECT textural features of normal salivary gland tissue that developed RT-induced xerostomia, another common complication of RT. In that study, the investigators used CECT textural features of salivary glands to create multivariable logistic regression models to predict RT-induced xerostomia. With regard to the utility of radiomic features for early detection of subclinical ORNJ specifically, we previously established a model to characterize temporal evolution of bone tissue prior to ORNJ development versus normal bone using CECT radiomic features change over time^26^. As a continuation of our efforts to characterize ORNJ using CECT radiomic features, the present study focused on the most important radiomic features that classify ORNJ (i.e., after diagnosis confirmation) compared with self-controlled normal mandibular tissue.

There are several limitations to our study. First, although the accuracy of our statistical model is highly satisfactory, it is notable that this accuracy is in the case of an already-established ORNJ versus control normal tissue that received minimal radiation. Therefore, these models may become less accurate in the case of pre-ORN mandibular subvolumes versus a neighboring control tissue that received a similar dose of radiation. To address this limitation, we will try to recruit more cases in our upcoming studies to maintain the same level of accuracy. We acknowledge the lack of a clinician-based accuracy benchmark for detecting ORN from imaging alone. Ongoing work is focused on establishing this benchmark, which will allow for a direct comparison to determine whether computerized methods offer a meaningful improvement over clinician evaluations. Given the potential for these computational approaches to outperform clinician assessments, this benchmark is essential for validation. Second, although all our cases were diagnosed by multiple radiologists, there is significant variability in the grade and severity of ORNJ. Finally, in this study we considered images that corresponded to the time of ORNJ clinical diagnosis, with the proportions of ORNJ grades skewed in favor of more advanced stages, when mandible damage was therefore very visible both to the clinical eye and on the CECT images. Expanding our study to include earlier images, prior to clinical diagnosis of ORNJ, will allow evaluation of subclinical tissue characteristics of ORNJ regions and subsequently enable earlier detection of mandibular damage.

Since we identified the most important radiomic features that differentiate ORNJ versus normal tissue, we plan to validate the utility of these features in the identification of at-risk areas well before the clinical or radiologic detection of ORNJ using a large-scale dataset. The next step would be to test the correlation between the temporal evolution of these radiomic features and the development of ORNJ. Successful prospective validation of an accurate statistical model to predict ORNJ would allow us to test the effect of early intervention via medical treatment and/or hyperbaric oxygen therapy to decrease the risk of ORNJ in susceptible patients.

Despite these limitations and the need for external validation of our results, this study yielded significant results, and we are hopeful that CECT texture analysis will likely also prove to be valid for early detection and/or prediction of ORNJ before clinical/radiologic diagnosis.

## Data Availability

All data produced are available online at https://figshare.com/s/728333f8ea94e555e77a.

## Acknowledgement

The authors would like to acknowledge the support from various funding sources that made this work possible. C.D.F. and S.Y.L. received related funding support from the NIH/NIDCR (U01DE032168/ R01DE025248). C.D.F. also receives infrastructure and salary support through the NIH/NCI MD Anderson Cancer Center Core Support Grant (CCSG) Image-Driven Biologically-informed Therapy (IDBT) program (P30CA016672-47), support from the NCI (R01CA257814) as well as additional unrelated salary/effort support from NIH institutes and philanthropic grants and infrastructure support from MD Anderson Cancer Center via the Charles and Daneen Stiefel Center for Head and Neck Cancer Oropharyngeal Cancer Research Program and the Program in Image-Guided Cancer Therapy. S.Y.L. is supported through the CCSG Head and Neck Program (P30CA016672-48). K.A.W. was supported by the Dr. John J. Kopchick Fellowship through The University of Texas MD Anderson UTHealth Graduate School of Biomedical Sciences, the American Legion Auxiliary Fellowship in Cancer Research, an NIH/NIDCR F31 fellowship (F31DE031502), and an Image-Guided Cancer Therapy (IGCT) T32 Training Program Fellowship (T32CA261856). Z.K.’s time was supported by a pre-doctoral fellowship from the Cancer Prevention Research Institute of Texas grant #RP210042. K.K.B. acknowledges support from the Image Guided Cancer Therapy Research Program at The University of Texas MD Anderson Cancer Center, which was partially funded by the National Institutes of Health/NCI under award number P30CA016672 and through a licensing agreement with RaySearch Laboratories AB. M.A.N. received funding from the National Institutes of Health/National Institute of Dental and Craniofacial Research (NIH/NIDCR) through grant 1R03DE033550-01. J.R. received salary support from the NIDCR Diversity Supplement Grant (3R01DE028290-02S1). L.V.V. received funding and salary support from KWF Dutch Cancer Society through a Young Investigator Grant (KWF-13529) and from NWO ZonMw through the VENI grant (NWO-09150162010173). A.S.R.M. received funding from NIDCR (U01DE032168, 1R01DE028290-01A1) and NCI (R01 CA258827). DF was supported by R01CA195524.

## Data availability statement

In accordance with the *Final NIH Policy for Data Management and Sharing*, NOT-OD-21-013, anonymized tabular analytic and NIFTI data that support the findings of this study are openly available in an NIH-supported generalist scientific data repository (https://figshare.com/s/728333f8ea94e555e77a) no later than the time of an associated peer—reviewed publication; while public data is embargoed pending peer review, the data is available upon request pre-peer-review through email to the corresponding author.

