## Supplementary Material for "Computed tomography radiomics-based cross-sectional detection of mandibular osteoradionecrosis in head and neck cancer survivors"

#### Appendix A. Clinical characteristics of the patients involved in the study.

| Characteristic | Value (N=150) |
|---|---|
| **Demographic Characteristics** | |
| Mean (Range) Age of Patients at the Time of the Contrast-Enhanced CT Study | 62.3 (27-82) |
| Sex | Male = 131 (87.3%),<br>Female: 19 (12.6%) |
| Race | White or Caucasian: 135 (90%),<br>Black or African American: 6 (4%),<br>Other: 9 (6%) |
| Smoking Status | Former: 85 (56.6%),<br>Never: 49 (32.6%),<br>Current: 16 (10.6%) |
| Mean Smoking Pack Years (SD) | 23.3 (29.5) |
| Alcohol Status | Yes: 53 (35.3%)<br>No: 93 (62%)<br>Former: 4 (2%) |
| Alcohol Levels * | None-Occasional-Moderate: 136 (90.7%)<br>Heavy: 14 (9.3%) |
| **Disease Chronology & Survivorship** | |
| RT End-$ORN_0$ Interval in Months | Mean: 32.6<br>SD: 41.6<br>Range: 0-282 |
| $ORN_0$-Mandibulectomy Interval in Months (n=69) | Mean: 19.6<br>SD: 20.6<br>Range: 1-90 |
| $ORN_0$-$ORN_1$ Interval in Months | Mean: 10<br>Range: 0-71 |
| $ORN_0$-Last Follow-up Interval in Years | Mean: 4.6<br>SD: 3.1<br>Range: 0-15 |
| Last Follow-up Status | Alive with ORN: 53 (35.3%)<br>No evidence of ORN: 41 (27.3%) |

| | |
|---|---|
| | Deceased: 56 (37.3%) |
| **ORN Characteristics** | |
| ORN Grade (Tsai et al. [18]) | Grade 1: 16 (10.7%),<br>Grade 2: 23 (15.3%),<br>Grade 3: 36 (24%),<br>Grade 4: 75 (50%) |
| ORN Laterality to Tumor Side | Ipsilateral: 127 (84.7%),<br>Contralateral: 23 (15.3%) |
| ORN Treatment Modalities | Mandibulectomy: 69 (46%)<br>Hyperbaric Oxygen: 79 (52.6%) |
| **RT Characteristics** | |
| RT Technique | IMRT: 103 (68.7%)<br>VMAT: 11 (7.3%)<br>IMPT: 9 (6%)<br>Conventional 3D Technique: 1 (0.7%)<br>Not Recorded: 26 (17.3%) |
| Pre-RT Dental Status | No Dental Intervention: 58 (38.7%)<br>Dental Extraction: 44 (29.3)<br>Edentulous: 12 (8%)<br>Not Recorded: 36 (24%) |
| Mean Radiation Dose (Range) | 66.6 (50-72) |
| Mean Number of Radiation Fractions (Range) | 32.3 (25-42) |
| The Patient had >1 Head and Neck RT Courses Prior to ORN (Yes/No) | No = 128 (85.3%)<br>Yes = 22 (14.7%) |
| Therapy Sequence | RT Only: 30 (20%)<br>CRT Only: 73 (48.7)<br>Induction CT followed by CRT: 39 (26%)<br>Induction CT followed by RT Only: 8 (5.3%) |
| **Tumor Characteristics** | |
| Tumor Site | Base of Tongue: 43 (28.7%),<br>Tonsil: 41 (27.3%),<br>Tongue: 21 (14%),<br>Floor of mouth: 8 (5.3%), |

| | |
|---|---|
| | Oropharynx: 6 (4%),<br>Parotid: 4 (2.7%),<br>Other Head and Neck Sites: 27 (18%) |
| Tumor Histology | Squamous Cell Carcinoma: 149 (99.3%),<br>Mucoepidermoid Carcinoma: 1 (0.7%) |
| T-Stage | T0: 1 (0.7%),<br>T1: 26 (17.3%),<br>T2: 44 (29.3%),<br>T3: 29 (19.3%),<br>T4: 38 (25.3%),<br>Unknown: 12 (8%) |
| N-Stage | N0: 34 (22.7%),<br>N1: 23 (15.3%),<br>N2: 80 (53.3%),<br>N3: 2 (1.3%),<br>Unknown: 11 (7.3%) |
| M-Stage | M0: 149 (99.3%),<br>M1: 1 (0.7%) |
| HPV Status | Positive: 55 (36.7%),<br>Negative: 9 (6%),<br>Not recorded: 86 (57.3%) |
| Tumor Recurrence (Yes/No) | Yes: 46 (30.7%),<br>No: 104 (69.3%) |

Abbreviations: $ORN_0$ is the date of the first clinical/radiologic detection of ORN. $ORN_1$ is the date at which the CT scan used in our study was taken. IMRT: Intensity-modulated radiotherapy. IMPT: Intensity-modulated proton therapy. VMAT: Volumetric modulated arc therapy. CT: Chemotherapy. CRT: Concurrent radiotherapy & chemotherapy.

* Heavy is defined as > 14 standard drinks per week for males or > 7 for females

** Last follow-up is considered the last clinical encounter with the patient in our institution.

#### Appendix B. Features selected through RFECV RF classifier.

| Rank | Feature | Normal Mandible | ORN | P-Value | Importance |
|---|---|---|---|---|---|
| 1 | wavelet-LLH_firstorder_Median | -1.10 (-2.87, 0.00) | 1.40 (0.52, 2.34) | < 0.01 | 0.062 |
| 2 | wavelet-LLH_firstorder_Mean | -1.95 (-4.30, -0.79) | 1.20 (-0.36, 3.05) | < 0.01 | 0.057 |
| 3 | exponential_gldm_DependenceEntropy | 6.87 (6.59, 7.12) | 6.40 (6.15, 6.62) | < 0.01 | 0.049 |
| 4 | wavelet-LLL_firstorder_90Percentile | 817.43 (773.61, 856.19) | 711.92 (659.39, 769.87) | < 0.01 | 0.044 |
| 5 | square_firstorder_Kurtosis | 2.03 (1.71, 2.75) | 3.50 (2.55, 4.68) | < 0.01 | 0.041 |
| 6 | square_firstorder_90Percentile | 177.60 (161.03, 190.71) | 135.85 (116.22, 157.83) | < 0.01 | 0.039 |
| 7 | square_glrlm_LongRunHighGrayLevelEmphasis | 1225.17 (919.73, 1539.25) | 694.74 (538.29, 875.75) | < 0.01 | 0.035 |
| 8 | wavelet-LLL_firstorder_InterquartileRange | 311.33 (268.92, 351.93) | 227.69 (181.54, 269.89) | < 0.01 | 0.032 |
| 9 | exponential_firstorder_Skewness | 0.98 (0.52, 1.49) | 1.93 (1.41, 2.54) | < 0.01 | 0.029 |
| 10 | exponential_firstorder_RootMeanSquared | 24.87 (19.76, 29.91) | 15.95 (13.31, 20.06) | < 0.01 | 0.028 |
| 11 | exponential_ngtdm_Contrast | 0.12 (0.07, 0.16) | 0.04 (0.03, 0.08) | < 0.01 | 0.027 |
| 12 | gradient_glcm_Idmn | 0.96 (0.95, 0.97) | 0.97 (0.97, 0.98) | < 0.01 | 0.026 |
| 13 | exponential_glcm_Autocorrelation | 57.78 (37.06, 81.57) | 22.07 (16.02, 34.62) | < 0.01 | 0.025 |
| 14 | exponential_glrlm_GrayLevelNonUniformityNormalized | 0.09 (0.08, 0.11) | 0.14 (0.12, 0.18) | < 0.01 | 0.025 |

| | | | | | |
|---|---|---|---|---|---|
| 15 | square_firstorder_Skewness | 0.44<br>(0.09, 0.86) | 1.16<br>(0.80, 1.49) | < 0.01 | 0.023 |
| 16 | exponential_firstorder_Kurtosis | 2.80<br>(2.10, 4.74) | 6.94<br>(4.13, 11.39) | < 0.01 | 0.021 |
| 17 | gradient_ngtdm_Contrast | 0.72<br>(0.50, 0.99) | 0.47<br>(0.36, 0.57) | < 0.01 | 0.019 |
| 18 | square_glrlm_RunEntropy | 6.02<br>(5.79, 6.14) | 5.70<br>(5.51, 5.85) | < 0.01 | 0.018 |
| 19 | wavelet-LLH_firstorder_90Percentile | 31.39<br>(27.55, 37.45) | 38.47<br>(32.91, 44.74) | < 0.01 | 0.017 |
| 20 | gradient_glcm_Contrast | 263.84<br>(167.73, 334.23) | 167.93<br>(115.00, 215.15) | < 0.01 | 0.016 |
| 21 | square_firstorder_Interquartile Range | 97.48<br>(76.20, 112.30) | 55.39<br>(39.96, 75.47) | < 0.01 | 0.015 |
| 22 | square_gldm_LargeDependenceHighGrayLevelEmphasis | 6611.11<br>(4109.85, 10103.91) | 3578.02<br>(2454.21, 4748.14) | < 0.01 | 0.014 |
| 23 | gradient_firstorder_Kurtosis | 2.76<br>(2.34, 3.51) | 3.25<br>(2.70, 3.78) | < 0.01 | 0.014 |
| 24 | exponential_glcm_JointEnergy | 0.05<br>(0.02, 0.10) | 0.14<br>(0.10, 0.21) | < 0.01 | 0.013 |
| 25 | square_glcm_JointAverage | 26.74<br>(22.33, 31.09) | 18.22<br>(15.78, 21.78) | < 0.01 | 0.013 |
| 26 | square_glcm_SumSquares | 301.68<br>(242.97, 381.81) | 183.56<br>(136.51, 250.74) | < 0.01 | 0.013 |
| 27 | wavelet-HLL_glcm_MCC | 0.58<br>(0.52, 0.65) | 0.55<br>(0.50, 0.59) | < 0.01 | 0.013 |
| 28 | gradient_glcm_DifferenceVariance | 110.70<br>(78.09, 143.84) | 74.90<br>(55.00, 100.18) | < 0.01 | 0.012 |
| 29 | exponential_glrlm_LongRunHighGrayLevelEmphasis | 112.34<br>(84.37, 148.09) | 69.26<br>(53.87, 91.57) | < 0.01 | 0.011 |
| 30 | wavelet-LLL_firstorder_Skewness | 0.14<br>(-0.23, 0.46) | 0.63<br>(0.35, 0.91) | < 0.01 | 0.011 |

| | | | | | |
|---|---|---|---|---|---|
| 31 | wavelet-HLL_firstorder_Median | -0.75<br>(-2.00, 0.35) | 0.09<br>(-0.55, 0.47) | < 0.01 | 0.010 |
| 32 | square_firstorder_Median | 73.87<br>(55.38, 103.63) | 44.23<br>(35.48, 56.59) | < 0.01 | 0.010 |
| 33 | wavelet-LLL_firstorder_Kurtosis | 1.96<br>(1.77, 2.43) | 2.69<br>(2.20, 3.39) | < 0.01 | 0.010 |
| 34 | exponential_glcm_MaximumProbability | 0.20<br>(0.11, 0.30) | 0.35<br>(0.28, 0.43) | < 0.01 | 0.009 |
| 35 | wavelet-LLH_firstorder_Range | 225.40<br>(190.42, 269.12) | 253.11<br>(215.54, 290.53) | < 0.01 | 0.009 |
| 36 | squareroot_firstorder_InterquartileRange | 94.05<br>(79.64, 107.28) | 74.94<br>(59.08, 87.19) | < 0.01 | 0.009 |
| 37 | wavelet-LLL_glszm_SizeZoneNonUniformityNormalized | 0.72<br>(0.69, 0.75) | 0.71<br>(0.68, 0.75) | 0.715 | 0.009 |
| 38 | wavelet-LLH_glszm_SizeZoneNonUniformityNormalized | 0.48<br>(0.45, 0.51) | 0.50<br>(0.46, 0.53) | < 0.01 | 0.009 |
| 39 | exponential_gldm_DependenceVariance | 37.96<br>(23.26, 52.92) | 57.36<br>(45.18, 71.97) | < 0.01 | 0.008 |
| 40 | wavelet-HLH_firstorder_Mean | 0.04<br>(-0.01, 0.12) | 0.00<br>(-0.07, 0.06) | < 0.01 | 0.008 |
| 41 | logarithm_firstorder_InterquartileRange | 48.06<br>(40.31, 57.05) | 41.90<br>(33.90, 48.22) | < 0.01 | 0.008 |
| 42 | logarithm_glcm_Imc1 | -0.21<br>(-0.24, -0.19) | -0.19<br>(-0.22, -0.17) | < 0.01 | 0.008 |
| 43 | squareroot_firstorder_Minimum | 193.13<br>(160.02, 212.86) | 119.76<br>(-54.24, 201.62) | < 0.01 | 0.008 |
| 44 | wavelet-LLL_firstorder_Minimum | 241.47<br>(185.86, 272.54) | 134.06<br>(-5.65, 247.62) | < 0.01 | 0.008 |
| 45 | logarithm_glszm_LargeAreaHighGrayLevelEmphasis | 165209.55<br>(63935.56, 582780.65) | 240066.10<br>(26617.54, 1869769.94) | 0.973 | 0.007 |

| | | | | | |
|---|---|---|---|---|---|
| 46 | wavelet-LLL_glcm_SumSquares | 3053.63 (2535.92, 3761.31) | 2336.29 (1711.19, 2976.32) | < 0.01 | 0.007 |
| 47 | original_glszm_SizeZoneNonUniformityNormalized | 0.55 (0.53, 0.58) | 0.56 (0.53, 0.59) | 0.612 | 0.007 |
| 48 | square_ngtdm_Contrast | 0.55 (0.41, 0.76) | 0.30 (0.22, 0.47) | < 0.01 | 0.007 |
| 49 | gradient_gldm_LargeDependenceHighGrayLevelEmphasis | 2066.87 (1733.78, 2428.37) | 1686.51 (1441.37, 1949.07) | < 0.01 | 0.006 |
| 50 | square_glcm_JointEnergy | 0.00 (0.00, 0.00) | 0.01 (0.00, 0.01) | < 0.01 | 0.006 |
| 51 | wavelet-LLH_firstorder_Maximum | 105.05 (86.84, 130.16) | 120.79 (98.98, 138.89) | < 0.01 | 0.006 |
| 52 | wavelet-HHH_glszm_SizeZoneNonUniformityNormalized | 0.45 (0.41, 0.47) | 0.45 (0.42, 0.47) | 0.927 | 0.006 |
| 53 | square_glcm_Imc1 | -0.21 (-0.25, -0.19) | -0.21 (-0.24, -0.17) | 0.1 | 0.006 |
| 54 | exponential_glcm_Imc2 | 0.86 (0.83, 0.89) | 0.82 (0.77, 0.85) | < 0.01 | 0.006 |
| 55 | wavelet-LLH_glszm_GrayLevelNonUniformityNormalized | 0.03 (0.03, 0.03) | 0.03 (0.02, 0.03) | 0.012 | 0.005 |
| 56 | wavelet-LHL_firstorder_Mean | -0.80 (-1.53, -0.39) | -0.73 (-1.27, -0.36) | 0.27 | 0.005 |
| 57 | gradient_glcm_ClusterShade | 13095.96 (7712.06, 18856.09) | 7795.60 (4734.61, 13476.16) | < 0.01 | 0.005 |
| 58 | logarithm_glcm_MaximumProbability | 0.03 (0.02, 0.04) | 0.02 (0.01, 0.03) | 0.015 | 0.005 |
| 59 | wavelet-HHH_glcm_Imc1 | -0.03 (-0.04, -0.02) | -0.02 (-0.03, -0.01) | < 0.01 | 0.005 |
| 60 | wavelet-LLL_glcm_Idmn | 0.98 (0.97, 0.99) | 0.99 (0.98, 0.99) | < 0.01 | 0.004 |
| 61 | gradient_glcm_ClusterProminence | 1453769.21 | 846369.68 | < 0.01 | 0.004 |

| | | | | | |
|---|---|---|---|---|---|
| | | (878408.89, 2255878.50) | (503966.68, 1431093.74) | | |
| 62 | exponential_glcm_MCC | 0.79 (0.74, 0.84) | 0.77 (0.71, 0.81) | < 0.01 | 0.004 |
| 63 | wavelet-LLH_glcm_DifferenceEntropy | 4.24 (4.01, 4.43) | 4.28 (4.11, 4.52) | 0.075 | 0.004 |
| 64 | logarithm_glcm_Idmn | 0.98 (0.97, 0.99) | 0.99 (0.98, 1.00) | < 0.01 | 0.004 |
| 65 | square_glszm_GrayLevelNonUniformityNormalized | 0.02 (0.02, 0.02) | 0.02 (0.02, 0.03) | < 0.01 | 0.003 |
| 66 | squareroot_ngtdm_Contrast | 0.47 (0.33, 0.66) | 0.32 (0.20, 0.51) | < 0.01 | 0.003 |
| 67 | squareroot_glcm_Imc1 | -0.21 (-0.25, -0.19) | -0.20 (-0.25, -0.17) | 0.12 | 0.002 |

### Appendix C. TRIPOD+AI checklist

Version: 11-January-2024

| Section/Topic | Item | Development / evaluation[1] | Checklist item | Reported on page |
|---|---|---|---|---|
| **TITLE** | | | | |
| *Title* | 1 | D;E | Identify the study as developing or evaluating the performance of a multivariable prediction model, the target population, and the outcome to be predicted | 1 |
| **ABSTRACT** | | | | |
| *Abstract* | 2 | D;E | See TRIPOD+AI for Abstracts checklist | |
| **INTRODUCTION** | | | | |
| *Background* | 3a | D;E | Explain the healthcare context (including whether diagnostic or prognostic) and rationale for developing or evaluating the prediction model, including references to existing models | 2 |
| | 3b | D;E | Describe the target population and the intended purpose of the prediction model in the context of the care pathway, including its intended users (e.g., healthcare professionals, patients, public) | 2/3 |
| | 3c | D;E | Describe any known health inequalities between sociodemographic groups | |
| *Objectives* | 4 | D;E | Specify the study objectives, including whether the study describes the development or validation of a prediction model (or both) | 3 |
| **METHODS** | | | | |
| *Data* | 5a | D;E | Describe the sources of data separately for the development and evaluation datasets (e.g., randomised trial, cohort, routine care or registry data), the rationale for using these data, and representativeness of the data | 3 |
| | 5b | D;E | Specify the dates of the collected participant data, including start and end of participant accrual; and, if applicable, end of follow-up | 3 |
| *Participants* | 6a | D;E | Specify key elements of the study setting (e.g., primary care, secondary care, general population) including the number and location of centres | 3 |
| | 6b | D;E | Describe the eligibility criteria for study participants | 3/4 |
| | 6c | D;E | Give details of any treatments received, and how they were handled during model development or evaluation, if relevant | |
| *Data preparation* | 7 | D;E | Describe any data pre-processing and quality checking, including whether this was similar across relevant sociodemographic groups | 4/5/6 |
| *Outcome* | 8a | D;E | Clearly define the outcome that is being predicted and the time horizon, including how and when assessed, the rationale for choosing this outcome, and whether the method of outcome assessment is consistent across sociodemographic groups | 3 |
| | 8b | D;E | If outcome assessment requires subjective interpretation, describe the qualifications and demographic characteristics of the outcome assessors | |
| | 8c | D;E | Report any actions to blind assessment of the outcome to be predicted | |
| *Predictors* | 9a | D | Describe the choice of initial predictors (e.g., literature, previous models, all available predictors) and any pre-selection of predictors before model building | 6 |
| | 9b | D;E | Clearly define all predictors, including how and when they were measured (and any actions to blind assessment of predictors for the outcome and other predictors) | 6 |
| | 9c | D;E | If predictor measurement requires subjective interpretation, describe the qualifications and demographic characteristics of the predictor assessors | |
| *Sample size* | 10 | D;E | Explain how the study size was arrived at (separately for development and evaluation), and justify that the study size was sufficient to answer the research question. Include details of any sample size calculation | 4/6 |
| *Missing data* | 11 | D;E | Describe how missing data were handled. Provide reasons for omitting any data | |
| *Analytical methods* | 12a | D | Describe how the data were used (e.g., for development and evaluation of model performance) in the analysis, including whether the data were partitioned, considering any sample size requirements | 6/7 |
| | 12b | D | Depending on the type of model, describe how predictors were handled in the analyses (functional form, rescaling, transformation, or any standardisation). | 6/7 |
| | 12c | D | Specify the type of model, rationale[2], all model-building steps, including any hyperparameter tuning, and method for internal validation | 6/7 |
| | 12d | D;E | Describe if and how any heterogeneity in estimates of model parameter values and model performance was handled and quantified across clusters (e.g., hospitals, countries). See TRIPOD-Cluster for additional considerations[3] | |
| | 12e | D;E | Specify all measures and plots used (and their rationale) to evaluate model performance (e.g., discrimination, calibration, clinical utility) and, if relevant, to compare multiple models | 7 |
| | 12f | E | Describe any model updating (e.g., recalibration) arising from the model evaluation, either overall or for particular sociodemographic groups or settings | |
| | 12g | E | For model evaluation, describe how the model predictions were calculated (e.g., formula, code, object, application programming interface) | 7 |
| *Class imbalance* | 13 | D;E | If class imbalance methods were used, state why and how this was done, and any subsequent methods to recalibrate the model or the model predictions | |
| *Fairness* | 14 | D;E | Describe any approaches that were used to address model fairness and their rationale | |
| *Model output* | 15 | D | Specify the output of the prediction model (e.g., probabilities, classification). Provide details and rationale for any classification and how the thresholds were identified | 7-10 |

[1] D=items relevant only to the development of a prediction model; E=items relating solely to the evaluation of a prediction model; D;E=items applicable to both the development and evaluation of a prediction model

[2] Separately for all model building approaches.

[3] TRIPOD-Cluster is a checklist of reporting recommendations for studies developing or validating models that explicitly account for clustering or explore heterogeneity in model performance (eg, at different hospitals or centres). Debray et al, BMJ 2023; 380: e071018 [DOI: 10.1136/bmj-2022-071018]

| | | | | |
|---|---|---|---|---|
| *Training versus evaluation* | 16 | D;E | Identify any differences between the development and evaluation data in healthcare setting, eligibility criteria, outcome, and predictors | |
| *Ethical approval* | 17 | D;E | Name the institutional research board or ethics committee that approved the study and describe the participant-informed consent or the ethics committee waiver of informed consent | 3 |
| **OPEN SCIENCE** | | | | |
| *Funding* | 18a | D;E | Give the source of funding and the role of the funders for the present study | 12/13 |
| *Conflicts of interest* | 18b | D;E | Declare any conflicts of interest and financial disclosures for all authors | 13 |
| *Protocol* | 18c | D;E | Indicate where the study protocol can be accessed or state that a protocol was not prepared | |
| *Registration* | 18d | D;E | Provide registration information for the study, including register name and registration number, or state that the study was not registered | |
| *Data sharing* | 18e | D;E | Provide details of the availability of the study data | 13 |
| *Code sharing* | 18f | D;E | Provide details of the availability of the analytical code[4] | |
| **PATIENT & PUBLIC INVOLVEMENT** | | | | |
| *Patient & Public Involvement* | 19 | D;E | Provide details of any patient and public involvement during the design, conduct, reporting, interpretation, or dissemination of the study or state no involvement. | |
| **RESULTS** | | | | |
| *Participants* | 20a | D;E | Describe the flow of participants through the study, including the number of participants with and without the outcome and, if applicable, a summary of the follow-up time. A diagram may be helpful. | 4 |
| | 20b | D;E | Report the characteristics overall and, where applicable, for each data source or setting, including the key dates, key predictors (including demographics), treatments received, sample size, number of outcome events, follow-up time, and amount of missing data. A table may be helpful. Report any differences across key demographic groups. | 7 |
| | 20c | E | For model evaluation, show a comparison with the development data of the distribution of important predictors (demographics, predictors, and outcome). | |
| *Model development* | 21 | D;E | Specify the number of participants and outcome events in each analysis (e.g., for model development, hyperparameter tuning, model evaluation) | 4 |
| *Model specification* | 22 | D | Provide details of the full prediction model (e.g., formula, code, object, application programming interface) to allow predictions in new individuals and to enable third-party evaluation and implementation, including any restrictions to access or re-use (e.g., freely available, proprietary)[5] | 6 |
| *Model performance* | 23a | D;E | Report model performance estimates with confidence intervals, including for any key subgroups (e.g., sociodemographic). Consider plots to aid presentation. | 8/9 |
| | 23b | D;E | If examined, report results of any heterogeneity in model performance across clusters. See TRIPOD Cluster for additional details[3]. | |
| *Model updating* | 24 | E | Report the results from any model updating, including the updated model and subsequent performance | |
| **DISCUSSION** | | | | |
| *Interpretation* | 25 | D;E | Give an overall interpretation of the main results, including issues of fairness in the context of the objectives and previous studies | 11/12 |
| *Limitations* | 26 | D;E | Discuss any limitations of the study (such as a non-representative sample, sample size, overfitting, missing data) and their effects on any biases, statistical uncertainty, and generalizability | 11/12 |
| *Usability of the model in the context of current care* | 27a | D | Describe how poor quality or unavailable input data (e.g., predictor values) should be assessed and handled when implementing the prediction model | 11/12 |
| | 27b | D | Specify whether users will be required to interact in the handling of the input data or use of the model, and what level of expertise is required of users | |
| | 27c | D;E | Discuss any next steps for future research, with a specific view to applicability and generalizability of the model | 11/12 |

From: Collins GS, Moons KGM, Dhiman P, et al. *BMJ* 2024;385:e078378. doi:10.1136/bmj-2023-078378

[4] This relates to the analysis code, for example, any data cleaning, feature engineering, model building, evaluation.
[5] This relates to the code to implement the model to get estimates of risk for a new individual.
